# Pulse pressure modifies the association between diastolic blood pressure and decrease in kidney function: The Japan Specific Health Checkups (J-SHC) Study

**DOI:** 10.1101/2023.09.21.23295933

**Authors:** Hiroyuki Tamaki, Masahiro Eriguchi, Hisako Yoshida, Takayuki Uemura, Hikari Tasaki, Riri Furuyama, Fumihiro Fukata, Masatoshi Nishimoto, Takaaki Kosugi, Masaru Matsui, Ken-ichi Samejima, Kunitoshi Iseki, Shouichi Fujimoto, Tsuneo Konta, Toshiki Moriyama, Kunihiro Yamagata, Ichiei Narita, Masato Kasahara, Yugo Shibagaki, Masahide Kondo, Koichi Asahi, Tsuyoshi Watanabe, Kazuhiko Tsuruya

## Abstract

**Background:** Unlike systolic blood pressure (SBP), the prognostic value of diastolic blood pressure (DBP) in kidney function has not been established. We hypothesized that pulse pressure (PP), which is associated with arteriosclerosis, would affect the prognostic value of DBP.

**Methods:** This longitudinal study used data from the Japan Specific Health Checkups Study conducted between 2008 and 2014. The participants were stratified into 3 PP subgroups (low-PP ≤39, normal-PP 40–59, and high-PP ≥60 mmHg). The exposures of interest were SBP and DBP, and the association between SBP/DBP and kidney outcomes (30% decline in the estimated glomerular filtration rate from baseline) was examined in each PP subgroup using a Cox proportional hazards model.

**Results:** Among 725,022 participants, 20,414 (2.8%) developed kidney outcomes during a median follow-up period of 34.6 months. Higher SBP was consistently associated with a higher incidence of kidney outcome in all PP subgroups. Although DBP had a positive linear association with the incidence of kidney outcome in low-and normal-PP subgroups, both lower (≤60 mmHg) and higher (≥101 mmHg) DBP were associated with a higher incidence of kidney outcome in high-PP subgroup with U-shaped curve. Hazard ratios (95% confidence intervals) of ≤60 mmHg (reference: 61-80 mmHg in normal-PP subgroup) and ≥101 mmHg were 1.26 (1.15–1.38) and 1.86 (1.62–2.14), respectively.

**Conclusion:** In this large population-based cohort, DBP was differently associated with kidney outcome by PP levels; lower DBP was significantly associated with a higher incidence of kidney outcome in high-PP subgroup but not in low-and normal-PP subgroups.

## Introduction

Chronic kidney disease (CKD) is characterized by a decreased glomerular filtration rate (GFR) and/or the presence of albuminuria. Its worldwide prevalence is increasing due to aging, smoking, and several comorbidities, including hypertension, diabetes, and obesity [^1-9^]. Since CKD is an independent risk factor for cardiovascular morbidity and mortality, preventing its development is of paramount importance [^10-12^]. Among these risk factors, hypertension is one of the most established risk factors contributing to the development of CKD.

Systolic blood pressure (SBP), diastolic blood pressure (DBP), and pulse pressure (PP) are generally used to evaluate blood pressure (BP). Previous reports have shown a positive linear association between SBP and the risk of kidney function decline [^9,13,14^]. Recently, a positive linear association between PP and the risk of kidney function decline has also been reported [^15,16^]. However, previous reports examining the relationship between DBP and kidney function have demonstrated inconsistent results, with some reporting positive linear associations [^6,13,17^] and others identifying no significant relationship [^18-20^]. Therefore, the prognostic value of DBP in kidney function remains unestablished.

The population with low DBP includes individuals with naturally low DBP as well as those with low DBP due to increased PP caused by diseases, including atherosclerosis and aortic valve regurgitation. We hypothesized that PP level could affect the prognostic value of DBP, and the difference in PP levels across previous studies might underlie the inconsistent relationship between DBP and kidney function. Therefore, we evaluated the association between DBP and kidney function within each stratified PP level. To the best of our knowledge, no study has examined the prognostic abilities of DBP and SBP for kidney function after stratification by PP within a large population-based cohort.

## Methods

### Study design

This longitudinal study was conducted based on databases derived from the Specific Health Checkup program in Japan conducted between 2008 and 2014 (The Japan Specific Health Checkups [J-SHC] study). The details of this cohort study and the program have been published previously [^21^]. Briefly, the Japanese government launched a program in 2008 to investigate early diagnosis of metabolic syndrome in the general population aged 40–74 years. Data were collected from the following 26 prefectures: Hokkaido, Yamagata, Miyagi, Fukushima, Tochigi, Ibaraki, Chiba, Saitama, Tokyo, Kanagawa, Niigata, Ishikawa, Fukui, Nagano, Gifu, Osaka, Hyogo, Okayama, Tokushima, Kochi, Fukuoka, Saga, Kumamoto, Oita, Miyazaki, and Okinawa. Participants with a single measurement of serum creatinine level, missing data for BP or serum creatinine level, and unreasonable outliers of baseline serum creatinine level (≥8 mg/dL or <0.3 mg/dL), as well as those having baseline blood pressure with DBP exceeding SBP, were excluded from this study.

### Variable

Data collection was based on a self-administered questionnaire, including past history of cardiovascular disease (CVD), stroke, current smoking status, alcohol consumption status, and medication usage. Current smoking was defined as having smoked more than 100 cigarettes or smoking for more than 6 months prior to the health check-up program. Height, body weight, and BP were measured by trained staff at each local medical institute. Office BP was measured using a standard sphygmomanometer or an automated device placed on the right arm, following a 5-min rest in the sitting position. PP was calculated as the difference between SBP and DBP. The exposures of interest were SBP, DBP, and PP. According to 2018 European Society of Cardiology/European Society of Hypertension Guidelines for the management of arterial hypertension, PP ≥60 mmHg was a risk factor for asymptomatic hypertension-mediated organ damage [^22^]. Therefore, we divided PP into three categories: low PP (≤39 mmHg), normal PP (40–59 mmHg) and high PP (≥60 mmHg). Urine and blood samples were collected after fasting overnight for 10 h. The urine dipstick test for proteinuria were expressed as −, ±, 1+, 2+, or 3+. All blood samples were assayed using an automatic clinical chemical analyzer within 24 h of collection at each local medical institute. Serum creatinine levels were measured using enzymatic methods. The estimated GFR (eGFR) was calculated using formulas developed for the Japanese population: eGFR (mL/min/1.73 m^2^) = 194 × serum creatinine (mg/dL)^−1.094^ × age (years)^−0.287^ (× 0.739 for women) [^23^].

The outcome of this study included the time leading to the first event of a 30% decline in eGFR from the baseline level. As described previously, a 30% decline in eGFR over 6 years serves as a predictor for end-stage kidney disease [^24^] and is established as a surrogate marker for chronic kidney disease (CKD) progression [^25-27^].

### Statistical analysis

The patients were categorized into three subgroups based on PP: low (≤39 mmHg), normal (40–59 mmHg), and high (≥60 mmHg) groups. When examining exposures as a categorical variable, DBP and SBP were categorized into 4 categories: ≤60, 61–80, 81–100, and ≥101 mmHg for DBP and ≤100, 101–130, 131–160, and ≥161 mmHg for SBP. Baseline data were expressed as means with standard deviations or numbers with percentages, as appropriate. Cox proportional hazard regression models were used to examine the association between baseline DBP or SBP and the time to the first event of 30% eGFR decline in each subgroup. The data were adjusted for sex, age, body mass index (BMI), history of CVD and stroke, current smoking status, antihypertensive and antidiabetic agents’ usage, HbA1c, and proteinuria as potential confounders. The nonlinear relationships between baseline BPs and kidney outcome were evaluated using restricted cubic spline (RCS) curves. Statistical significance was set at a P-value <0.05. All statistical analyses were performed using R version 4.1.2 (R Foundation, Vienna, Austria).

### Ethics approval and consent to participate

All procedures involving human participants were conducted in accordance with the ethical standards of the institutional and/or national research committee under whose jurisdiction the studies were conducted (Fukushima Medical University; IRB Approval Number #1485, #2771) and principles outlined in the 1964 Helsinki Declaration and its later amendments. This study was conducted in accordance with the Ethical Guidelines for Medical and Health Research Involving Human Subjects enacted by the Ministry of Health, Labour, and Welfare of Japan ta[http://www.mhlw.go.jp/file/06-Seisakujouhou-10600000nDaijinkanboukouseikagakuka/0000069410.pdf].

## Results

### Baseline characteristics of the study cohort

Among the 933,488 patients, 208,466 were excluded based on the exclusion criteria, and the remaining 725,022 were included in this study (Figure 1). Baseline characteristics according to DBP and SBP are listed in Table 1. Those with higher DBP exhibited lower likelihood of being female; exhibited higher BMI, SBP, PP, and serum creatinine levels; and included a larger proportion of patients receiving antihypertensive agents and experiencing proteinuria (Table 1). Higher SBP was associated with lower likelihood of being female; older age; higher BMI, DBP, PP, and HbA1c levels; and higher prevalence of patients receiving antihypertensive agents and experiencing proteinuria (Table 1). Supplemental Tables S1–S6 present baseline characteristics according to DBP and SBP and further stratified based on PP categories. The association between the baseline characteristics and DBP in each PP group was similar compared to their association before PP stratification (Supplemental Tables S1– S3). However, higher DBP was associated with younger age only in the normal (40–59 mmHg) and high PP (≥60 mmHg) groups (Supplemental Tables S2 and S3). Baseline characteristics according to SBP in each PP group were similar to those before PP stratification (Supplemental Tables S4–S6).

**Figure 1.**
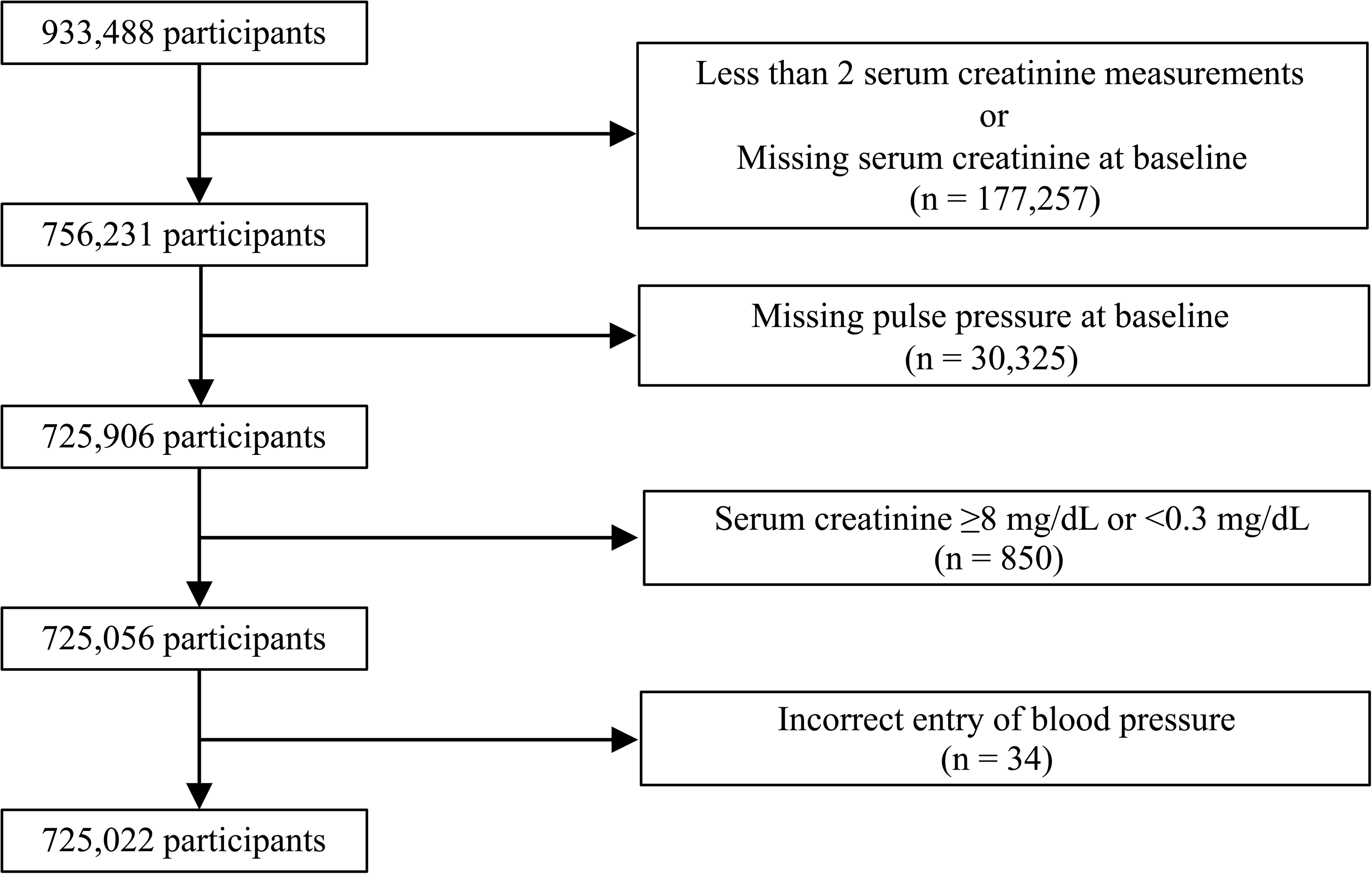
Flowchart of the study participants

**Table 1.**
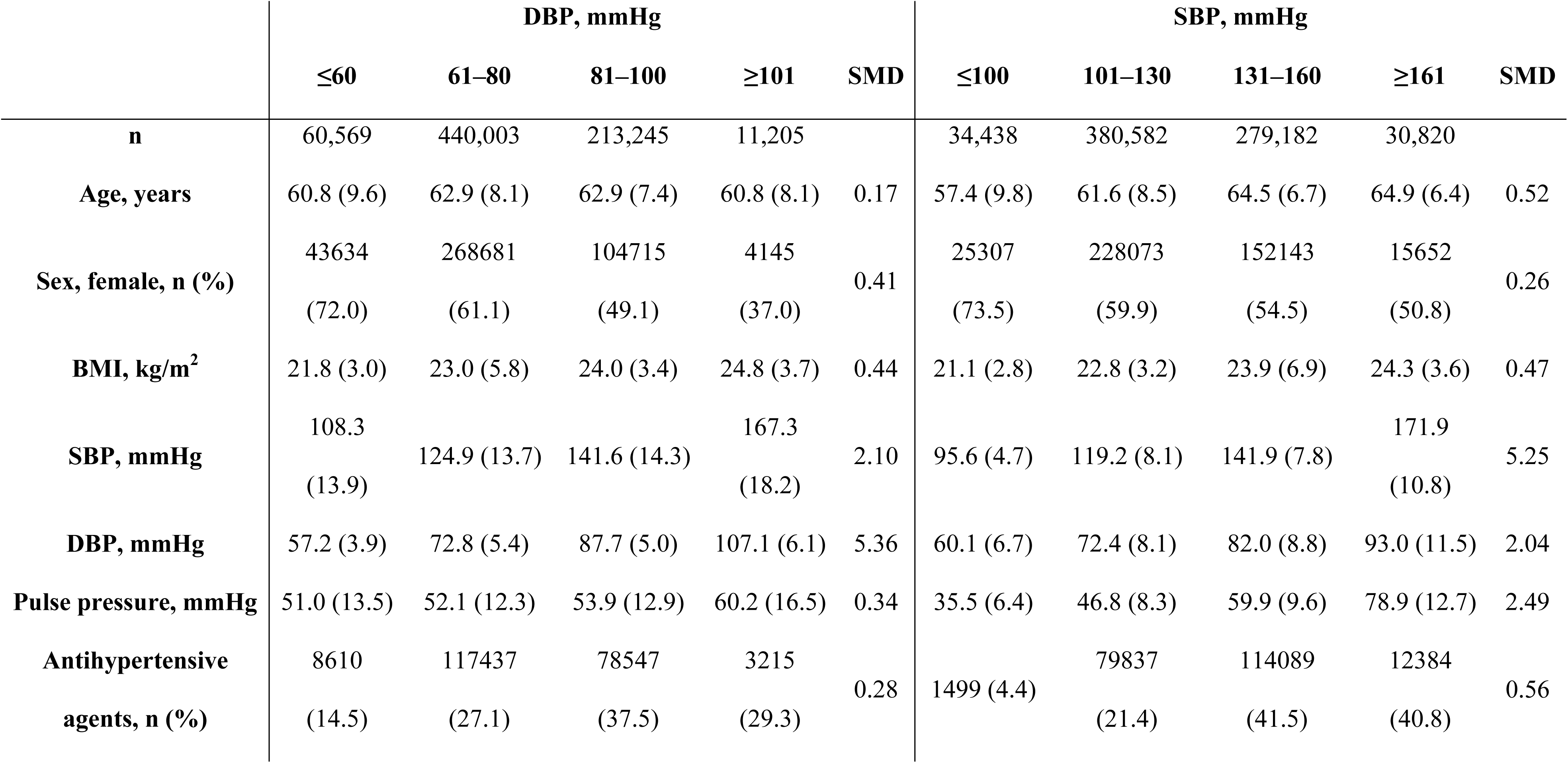
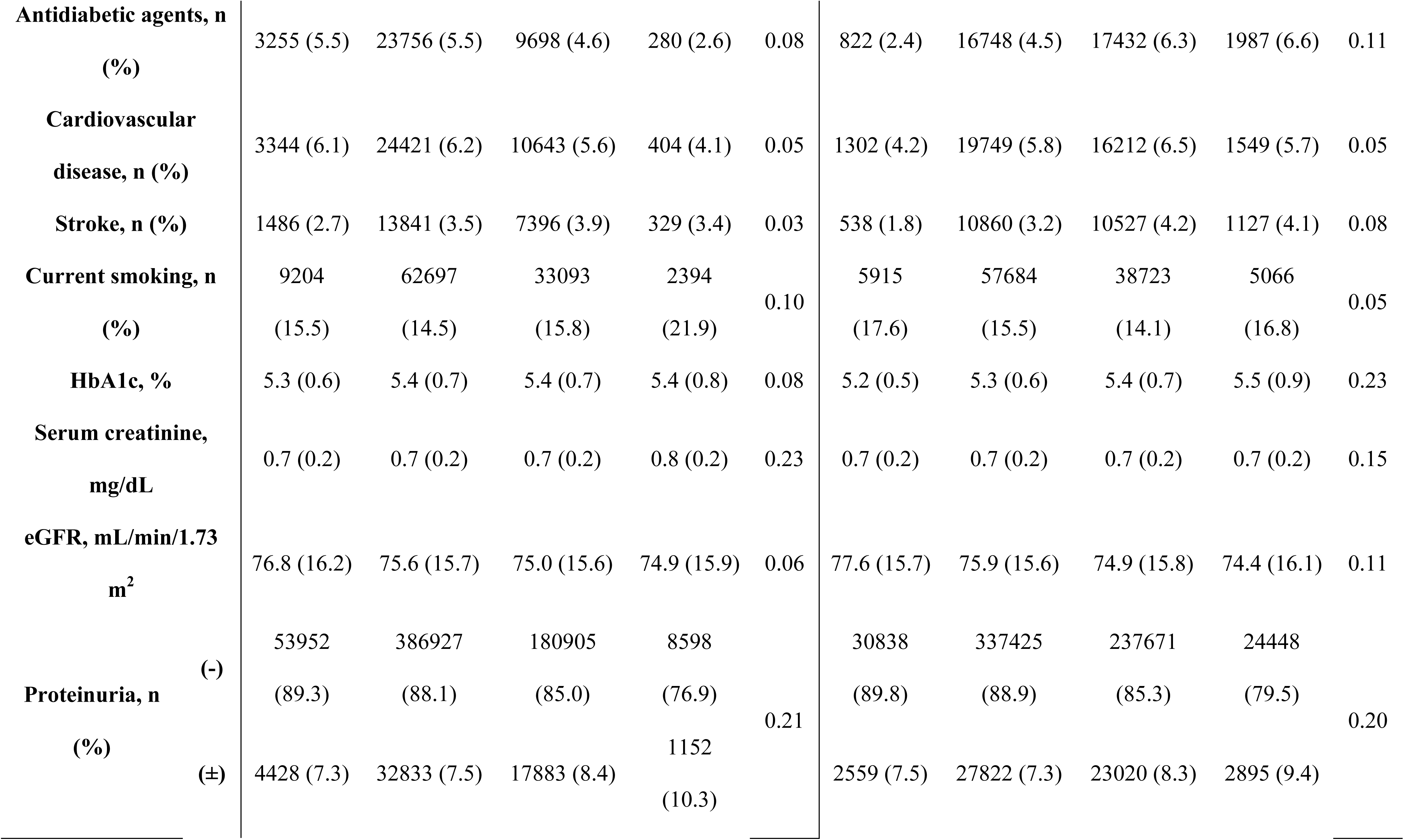
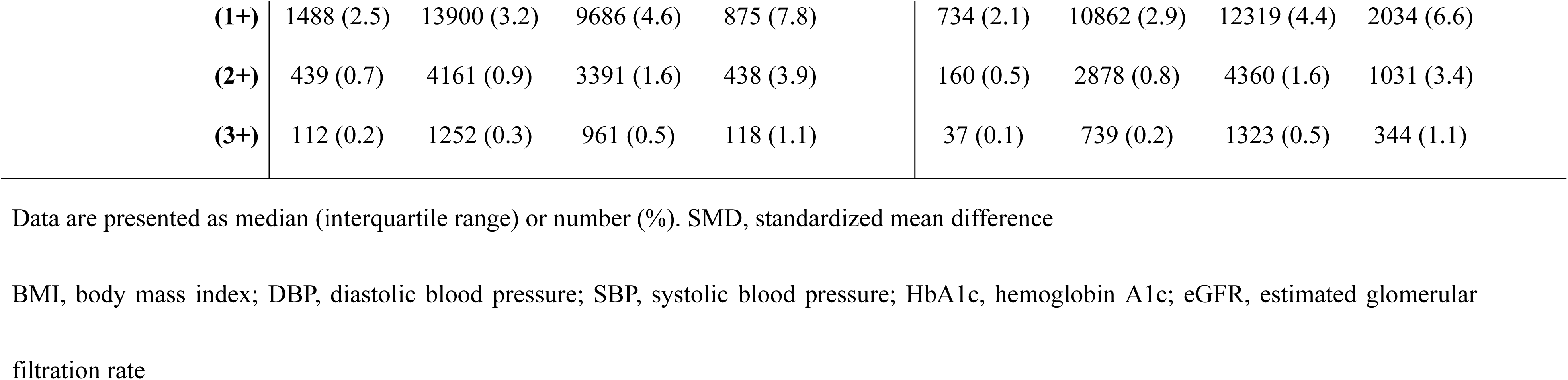
Baseline characteristics according to diastolic blood pressure and systolic blood pressure.

### Association of DBP with kidney outcome based on different PP levels

During a median follow-up of 34.6 months, 20,414 of 725,022 patients exhibited a 30% decline in eGFR from the baseline levels. Using Cox hazard models adjusted for three progressive sets of potential confounders, the hazard ratios (HRs) of DBP categories for kidney outcome were demonstrated within each PP subgroup, as presented in Table 2 (reference: participants with DBP 61–80 mmHg in the normal PP subgroup [40–59 mmHg]). The fully adjusted HRs of DBP categories in each PP subgroup are displayed in Figure 2A. Furthermore, we examined the risk of DBP using RCS within each PP subgroup. (Figure 3A, 3B) As depicted in Figure 2A, higher PP categories were associated with a higher incidence of kidney outcome within each DBP category. Linear positive associations between DBP and the incidence of kidney outcome were observed in the low and normal PP subgroups (Figure 2A, 3B). In the low PP subgroup, the fully adjusted HRs (95% confidence intervals [CIs]) of the highest (≥101 mmHg) and lowest (≤60 mmHg) DBP categories were 1.21 (0.78–1.88) and 0.96 (0.83–1.12), respectively (Figure 2A and Table 2). Similarly, in the normal PP subgroup, the fully adjusted HRs (95% CIs) of the highest (≥101 mmHg) and lowest (≤60 mmHg) DBP categories were 1.45 (1.22–1.72) and 0.93 (0.86–1.01), respectively (Figure 2A and Table 2). In contrast, both lower and higher DBPs were associated with a higher incidence of kidney outcome in the high-PP subgroup (Figure 2A, 0013B). The fully adjusted HRs (95% CIs) of the highest (≥101 mmHg) and lowest (61–80 mmHg) DBP category were 1.86 (1.62–2.14) and 1.11 (1.07–1.16), respectively (Figure 2A and Table 2). As illustrated in Figure 3A and 3B, the RCS curves demonstrated a U-shaped association between DBP and the incidence of kidney outcome in the overall cohort and high PP subgroup. However, a positive linear association between DBP and the incidence of kidney outcome was observed in the low and normal PP subgroups (Figure 3B).

**Figure 2.**
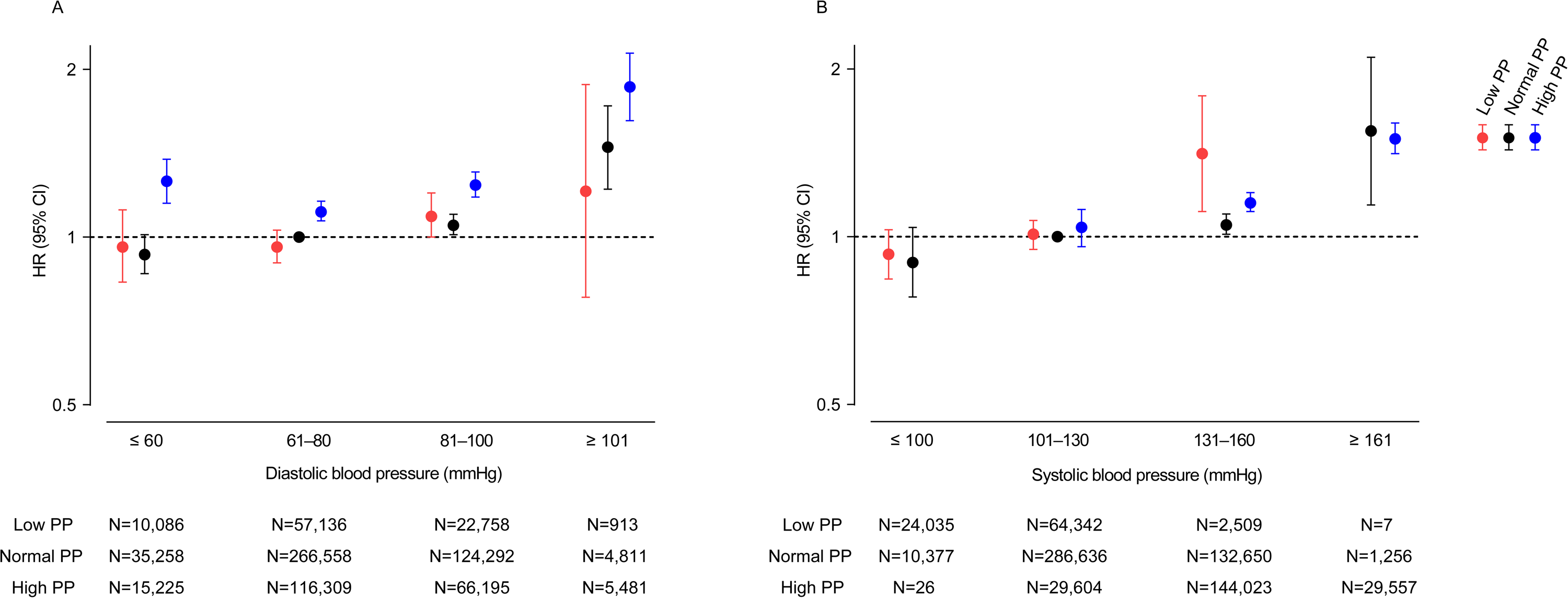
Fully adjusted HRs (95% CIs) for 30% eGFR reduction from baseline level based on DBP and PP **(A)**, and SBP and PP **(B)**. HRs and 95% CIs are shown as closed circles and vertical lines, respectively, in red (low PP), black (normal PP), and blue (high PP). HR, hazard ratio; CI, confidence interval; PP, pulse pressure; DBP, diastolic blood pressure; SBP, systolic blood pressure; eGFR, estimated glomerular filtration rate

**Figure 3.**
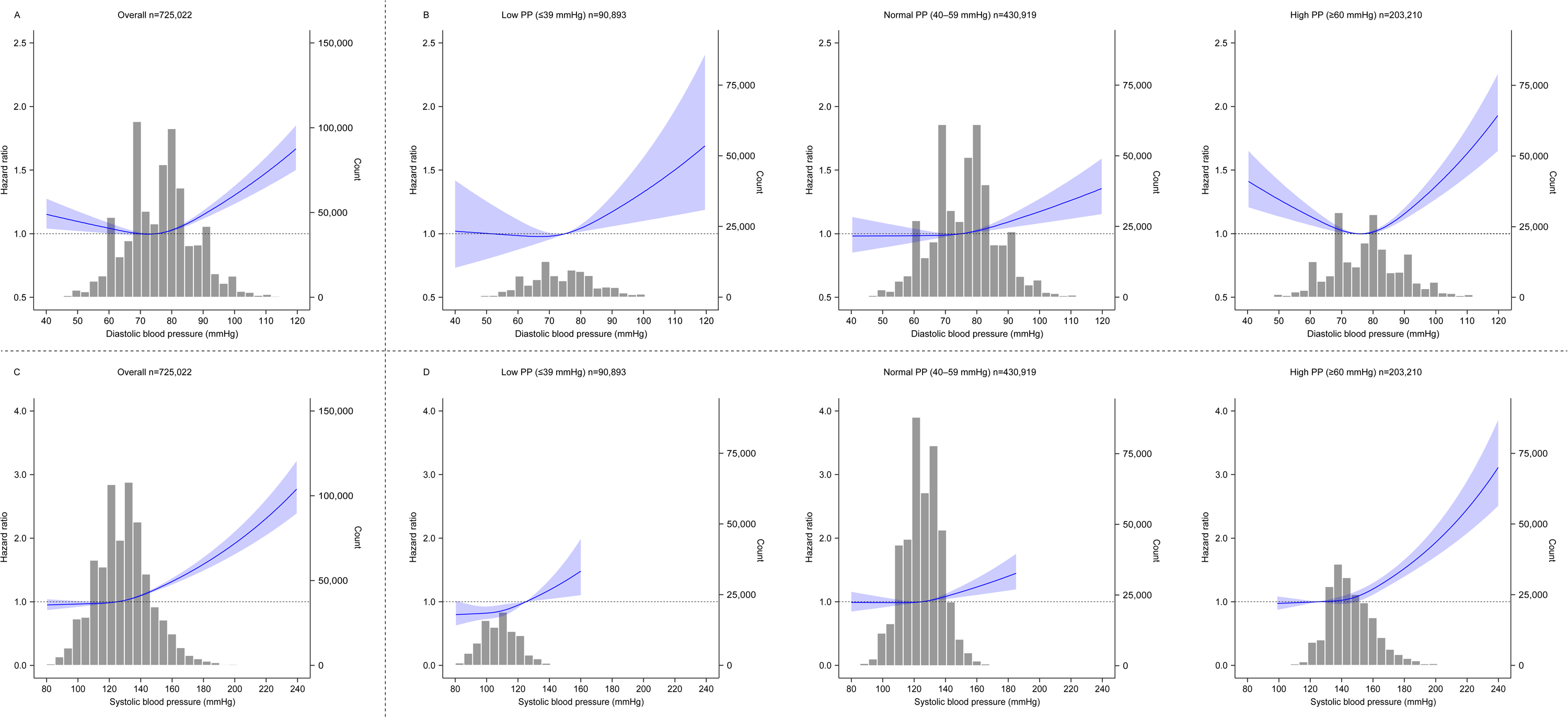
The restricted spline curves showing fully adjusted HRs (solid lines) and 95% CIs (shaped areas) for the relationships between 30% eGFR reduction from baseline and baseline diastolic **(A, B)** and systolic **(C, D)** blood pressures in all participants **(A, C)** and those in each PP category **(B, D)** HR, hazard ratio; CI, confidence interval; PP, pulse pressure; eGFR, estimated glomerular filtration rate

**Table 2.**
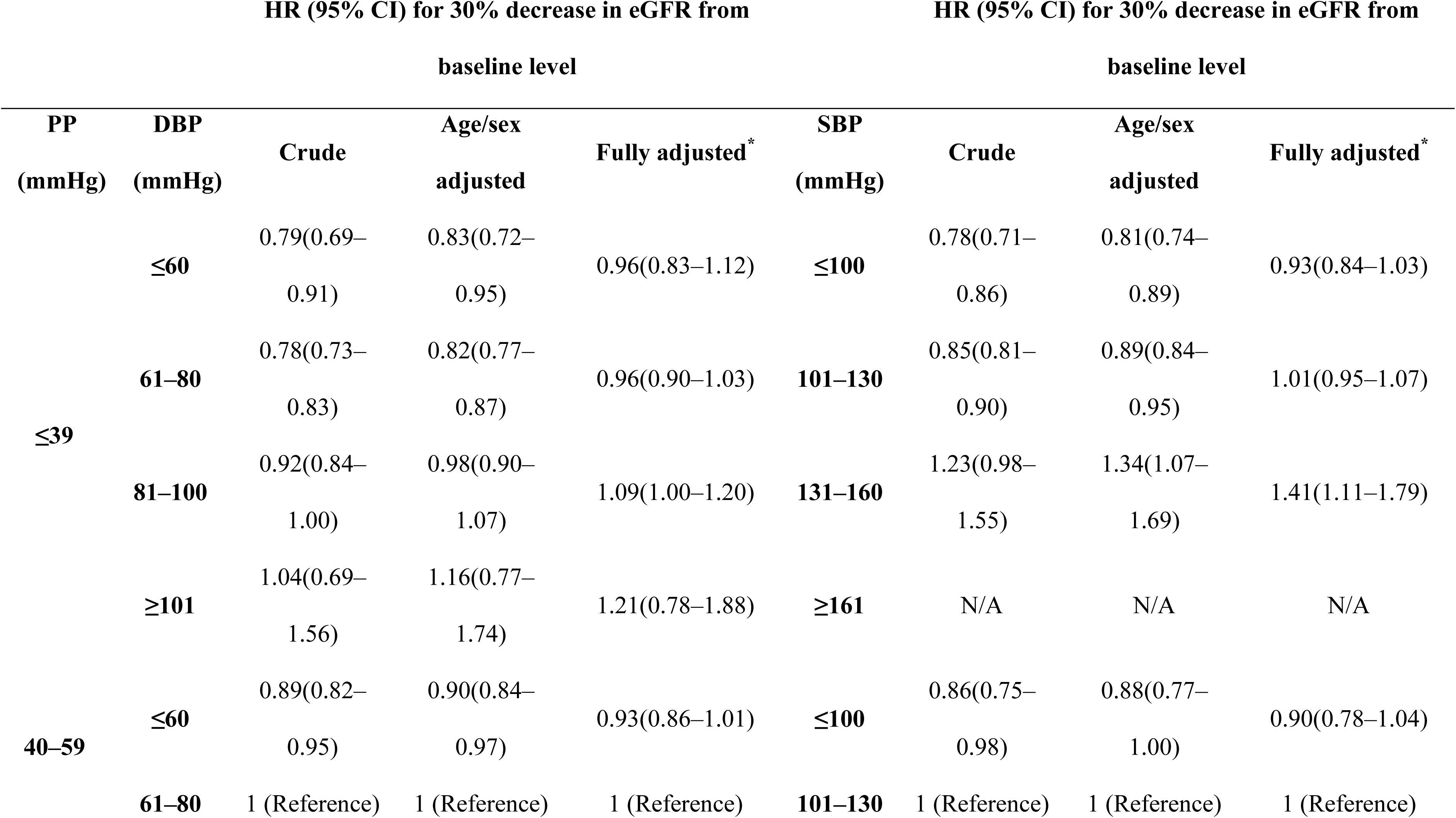
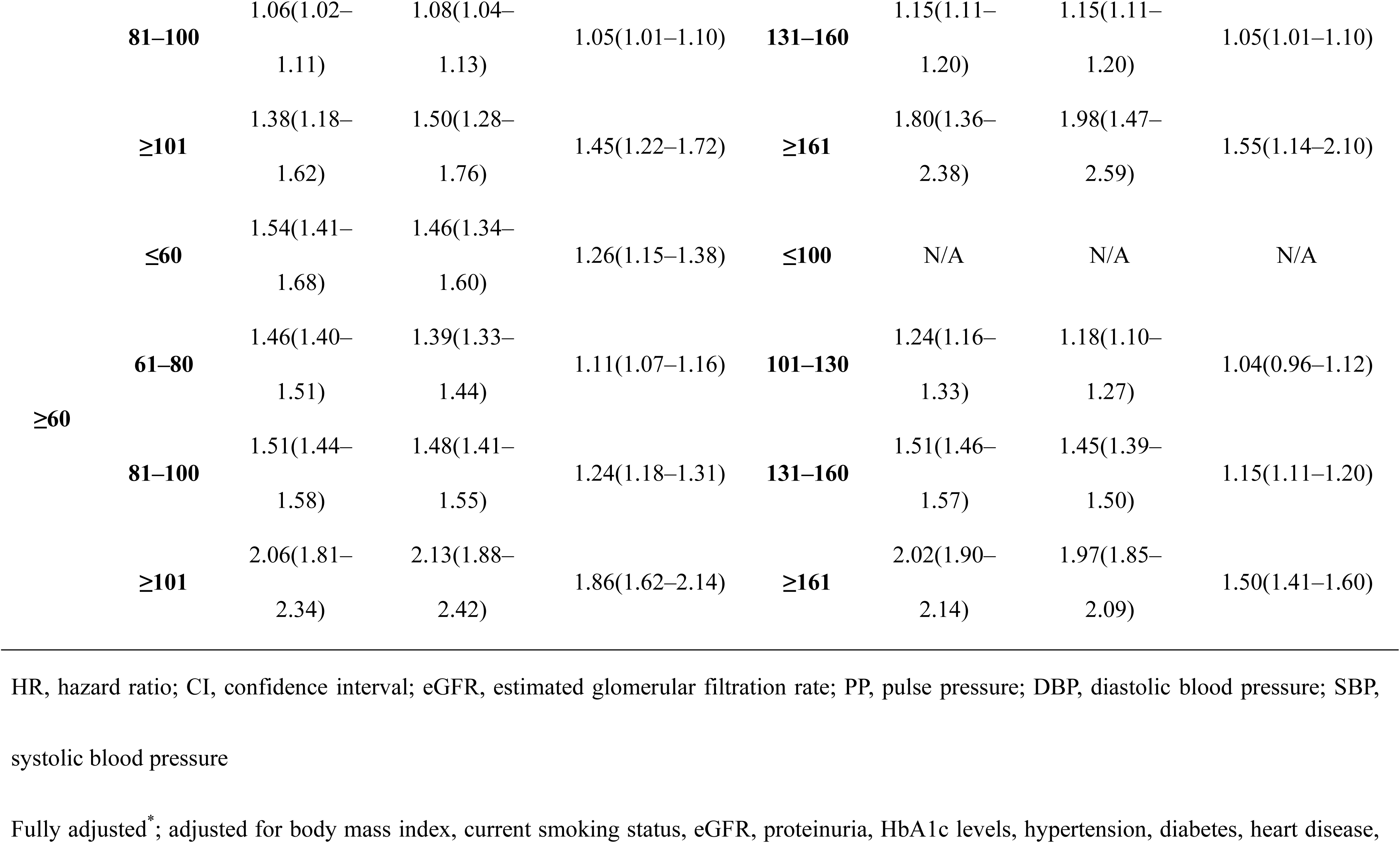
HRs (95% CIs) for 30% eGFR reduction from baseline according to DBP and SBP in stratified PP groups.

### Association of SBP with kidney outcome based on different PP levels

Using Cox hazard models adjusted for three progressive sets of potential confounders, the HRs of SBP categories for kidney outcome were demonstrated within each PP subgroup, as presented in Table 2 (reference: participants with SBP 101–130 mmHg in the normal PP subgroup [40–59 mmHg]). The fully adjusted HRs of SBP categories in each PP subgroup are displayed in Figure 2B. Furthermore, we examined the risk of SBP using RCS within each PP subgroup (Figure 3C, 3D). In contrast to the results observed for each DBP category, higher PP values were not associated with a higher incidence of kidney outcome in each SBP category (Figure 2B). Linear positive associations between SBP and the incidence of kidney outcome were observed within all PP subgroups (Figure 2B, 3D). In the low PP subgroup, the fully adjusted HRs (95% CIs) of the highest (≥131 mmHg) and lowest (≤100 mmHg) SBP categories were 1.41 (1.11–1.79) and 0.93 (0.84–1.03), respectively (Figure 2B and Table 2). In the normal PP subgroup, the fully adjusted HRs (95% CIs) of the highest (≥161 mmHg) and lowest (≤100 mmHg) SBP categories were 1.55 (1.14–2.10) and 0.90 (0.78–1.04), respectively (Figure 2B and Table 2). In high PP subgroup, the fully adjusted HRs (95% CIs) of the highest (≥161 mmHg) and lowest (≤130 mmHg) SBP categories were 1.50 (1.41–1.60) and 1.04 (0.96–1.12), respectively (Figure 2B and Table 2). As illustrated in Figures 3C and 3D, the RCS curves demonstrated positive linear associations between SBP and the incidence of kidney outcome in the overall cohort and all PP subgroups.

## Discussion

The present study demonstrated a positive linear association between SBP and poor kidney outcome and a U-shaped association between DBP and poor kidney outcome in a large general population-based cohort. Upon stratifying PP levels, the positive linear association with SBP was consistent within all PP subgroups. In contrast, the positive linear association of DBP with poor kidney outcome was observed in the low (≤39 mmHg) and normal PP (40– 59 mmHg) subgroups, whereas a U-shaped association of DBP was observed only in the high PP subgroup (≥60 mmHg). Furthermore, we observed that higher PP levels were associated with poor kidney outcome, regardless of DBP; however, this association was not evident when participants were categorized based on SBP.

As confirmed by previous studies [^9,13,14^], SBP is a strong predictor of kidney outcomes, and notably, our study demonstrated that SBP was a reliable predictor, regardless of the PP levels. In contrast, the prognostic value of DBP for kidney outcomes exhibited variability depending on the PP levels.

Previous studies investigating the association between DBP and kidney function have reported inconsistent results [^6,13,17-20^]. Some studies have reported linear positive association between DBP and poor kidney outcomes [^6,13,17^], while others have failed to establish a significant association between them [^18-20^]. The potential influence of PP variation among the patients in each previous study might have influenced this disparity. Indeed, the mean PP levels in studies that reported a significant association between DBP and kidney outcomes were within the normal range (46–52 mmHg) [^6,13,17^], but those in studies that failed to establish a significant association revealed high PP levels (65–93 mmHg) [^18-20^]. These results are consistent with our findings that a significant linear association between DBP and poor kidney outcome was observed in participants with low to normal PP levels but not in participants with high PP levels.

Although the mechanism underlying these findings remains unknown, we initially hypothesized that the increased risk of poor kidney outcome due to low DBP in participants with high PP could be attributed to increased arterial stiffness, which is associated with low DBP and high PP levels [^28,29^]. Contrary to our expectations, the participants in our study exhibited a weak correlation between DBP and PP (R = 0.11, Supplemental Figure S1), and a very strong correlation between SBP and PP (R = 0.79, Supplemental Figure S2), indicating that increased PP levels due to arteriosclerosis contributed to increased SBP rather than decreased DBP. These outcomes suggested that the mechanism linking poor kidney outcome to decreased DBP is not related to arteriosclerosis (arteriosclerosis-induced increased PP levels are not associated with decreased DBP). A U-shaped association between DBP and poor prognosis, as observed in the present study, has also been reported in a previous study describing the association between CVD and cardiovascular death and DBP [^30,31^]. This finding is supported by the fact that coronary artery perfusion occurs mainly during diastole and is, therefore, strongly influenced by decreased DBP [^32^]. Conversely, a previous report described that the effect of DBP on renal hemodynamics is relatively small [^33^]. Notably, older individuals and those with advanced arterial stiffness are highly susceptible to organ hypoperfusion due to impaired hemodynamic autoregulatory mechanisms [^33,34^]. The baseline characteristics in this study indicated that participants with high PP level were older, had a higher usage of antihypertensive and antidiabetic agents, and had a higher prevalence of a history of CVD than those of individuals with low-to-normal PP levels. The impaired hemodynamic autoregulatory system caused by arteriosclerosis (older age and increased comorbidities) could be one of the mechanisms underlying low-DBP-susceptible kidney ischemic injury.

As previously demonstrated, a higher PP level is associated with a higher incidence of kidney outcome [^15,16^]. We also validated this association when categorizing participants based on their DBP levels. Unexpectedly, this association was not significant when participants were categorized based on their SBP levels. In the present study, a strong association between PP and SBP (Supplemental Figure S2) was observed, and this association was also observed within the each categorized DBP level. Therefore, it is reasonable that higher PP levels as well as SBP levels were both associated with poor kidney outcomes when stratified based on DBP levels (within the same DBP level). In contrast, higher PP levels mean lower DBP levels within a narrow range of SBP (when stratified based on SBP levels), suggesting that higher PP levels (lower DBP levels) were not clearly associated with increased risk of kidney outcomes within a restricted range of SBP in the present study.

This study had several limitations. First, because this was an observational study, we could not determine a causal relationship between DBP and decreased kidney function. Potential confounding factors could have led to an increase or a decrease in DBP. Second, we could not obtain data on thyroid or valvular heart disease, which affect DBP and PP. Therefore, we were unable to determine the effects of these diseases on DBP or kidney function. Third, we estimated the GFR using creatinine-based equations. Therefore, the estimated kidney function of participants with poor nutritional status or lean body mass may not be accurate.

In conclusion, in this large population-based cohort, higher SBP and DBP were consistently associated with a higher incidence of kidney outcomes, regardless of PP levels; however, lower DBP was also significantly associated with a higher incidence of kidney outcome in participants with high PP levels, but not in those with low and normal PP levels. Attention should be paid not only to patients with high DBP but also to those with low DBP and high PP levels, as they could be at risk of adverse kidney outcomes.

## Data Availability

Due to the nature of this research, participants of this study did not agree for their data to be shared publicly, so supporting data is not available.

## Non-standard Abbreviations and Acronyms

BMI: body mass index
BP: blood pressure
CKD: chronic kidney disease
CI: confidence interval
CVD: cardiovascular disease
DBP: diastolic blood pressure
eGFR: estimated glomerular filtration rate
HR: hazard ratio
PP: pulse pressure
RCS: restricted cubic spline
SBP: systolic blood pressure

## Acknowledgments

The authors acknowledge the contributions of the staff members who collected the data and instructed the participants on metabolic syndrome at screening centers in the following regions: Hokkaido, Yamagata, Miyagi, Fukushima, Tochigi, Ibaraki, Chiba, Saitama, Tokyo, Kanagawa, Niigata, Ishikawa, Fukui, Nagano, Gifu, Osaka, Hyogo, Okayama, Tokushima, Kochi, Fukuoka, Saga, Kumamoto, Oita, Miyazaki, and Okinawa.

## Supplemental Material

Tables S1, S2, S3, S4, S5, S6

Figure S1, S2

## Author contributions

Research idea and study design: HTam, ME, TU, HTas, RF, FF, MN, TaK, MM, KS, KT; data acquisition: HY, KI, SF, TsK, TM, KY, IN, MKa, YS, MKo, KA, TW, KT; data analysis/interpretation: HTam, ME, KT; statistical analysis: HTam, ME; supervision or mentorship: ME, KT. Each author contributed valuable intellectual content throughout manuscript drafting or revision, and accepted accountability for the overall work by ensuring that questions pertaining to the accuracy or integrity of any portion of the work were appropriately investigated and resolved.

